# Rapid detection of myeloid neoplasm fusions using Single Molecule Long-Read Sequencing

**DOI:** 10.1101/2022.06.16.22276469

**Authors:** Olga Sala-Torra, Shishir Reddy, Ling-Hong Hung, Lan Beppu, David Wu, Jerald Radich, Ka Yee Yeung, Cecilia CS Yeung

## Abstract

Recurrent gene fusions are common drivers of disease pathophysiology in leukemias. Identification of these structural variants helps stratify disease by risk and assists with therapy choice. Current fusion detection methods require long turnaround time (7-10 days) or advance knowledge of the genes involved in the fusions. To address the need for rapid identification of clinically actionable fusion genes in heme malignancies without *a-priori* knowledge of the genes, we describe a long-read sequencing DNA assay designed with CRISPR guides to select and enrich for recurrent leukemia fusion genes. By applying rapid sequencing technology based on nanopores, we sequenced long pieces of genomic DNA and successfully detected fusion genes in cell lines and primary specimens (e.g., *BCR-ABL1, PML-RARA, CBFB-MYH11, KMT2A-AF4*) using cloud-based bioinformatics workflows with novel custom fusion finder software. We detected fusion genes in 100% of cell lines with the expected breakpoints and confirmed the presence or absence of a recurrent fusion gene in 12 of 14 patient cases. With our optimized assay and cloud-based bioinformatics workflow, these assays and analyses could be performed in under 8 hours.

**Key points:**

1. We describe a CRISPR-Cas9 enrichment Nanopore sequencing assay with streamlined bioinformatics that outperforms other fusion detectors.
2. We successfully detected both fusion genes and specific breakpoints in CML, APL, and AML in under 8 hours in 80% of patients.

**Visual Abstract (Figure 1):**

1. We successfully detected fusion genes in hematological malignancies with a fast and efficient long-read sequencing workflow in under 8 hours. The method makes the genomic characterization of *BCR-ABL1* DNA breakpoint in patients quick and simple, which potentiates design of patient specific primers for personalized monitoring MRD assays.
2. Our assay is based on a CRISPR-Cas9 non-amplification enrichment library preparation strategy and uses Nanopore sequencing single stranded genomic DNA coupled with streamlined bioinformatic workflow containing a novel fusion detector software which outperforms current fusion detection software.

## Introduction

Current classification of myeloid malignancies is largely based on the molecular and genetic aberrations ^1,2^. Recurrent gene rearrangements are present in 30-40% of acute myeloid leukemias (AML), and well described driver fusions that in some cases suffice to diagnose leukemias with *PML-RARA, RUNX1-RUNX1T1*, etc. even when the blast percentage is below 20%^1^. Recurrent fusion genes confer certain clinical and biological characteristics as drivers of leukemogenesis, and their identification assists in prognosis stratification and inform treatment decisions. Identification of driver fusion genes is especially relevant when targeted therapies are available. Examples include tyrosine kinase inhibitors (TKI) in chronic myelogenous leukemia (CML)^3^ that bind to the kinase domain in ABL1 deregulated as a consequence of the fusion with BCR, and differentiation therapy in acute promyelocytic leukemia (APL) in which identification of the promyelocytic leukemia-retinoic acid receptor alpha (*PML-RARA*) fusion confirms the diagnosis and indicates that the patient will likely respond to treatment with all-trans retinoic acid (ATRA) therapy, a nontoxic and highly effective treatment^4^ that achieves 90% long term survival rates when combined with anthracyclines and/or arsenic trioxide.^5^

Fusion genes can be detected through several techniques that include cytogenetics techniques such as fluorescence *in situ* hybridization (FISH), and molecular testing such as polymerase chain reaction (PCR) and next generation sequencing (NGS), which are the clinical gold standard. Clinical molecular assays for *BCR-ABL1* and *PML-RARA* primarily target RNA transcripts by RT-PCR because of the greater abundance of fusion gene transcripts compared to the DNA copies per cell, and because of the large variability in the genomic sequence of the fusion that can encompass large intronic region making routine DNA PCR impossible (e.g., *BCR-ABL*).

Typical NGS reads are 150 to 250 bps in length which are not sufficiently long enough to extend over DNA fragments that are adjoined because of a large genomic aberration thus hindering both alignment and detection of large indels and other structural abnormalities. NGS is limited by short read assembly mis-mapping, and amplification strategies instill imperfect quantitation of the variant allele frequencies. The diversity of aberrations in myeloid neoplasms include large genomic aberrations, including insertions and deletions, loss of heterozygosity, single nucleotide polymorphism, mutations in homopolymer rich regions and highly repetitive regions such as internal tandem duplications, that are difficult to detect by NGS and require different assays in molecular and cytogenetics labs additionally to those used to confirm fusion genes. Hence, currently a multiple assay approach is used to obtain a complete diagnostic molecular picture in myeloid malignancies.

The advancement of long-read sequencing technologies has enabled the sequencing of continuous single DNA or RNA molecules up to tens to hundreds of kilobases (kb) long.^6^ Ongoing improvements in Nanopore sequencing accuracy have reduced error rates to less than 5%,^7^ but remain higher than those for Illumina and Ion Torrent, which are used frequently in clinical laboratories.^8^ This technology has already made an impact on the understanding of the pathobiology of various diseases,^9,10^ and its impact will increase as the quality of sequencing improves and becomes more accurate.^7,11^ Addition of CRISPR-Cas9 for targeted enrichment concentrates the regions of interests prior to sequencing by Nanopore^12^ without requiring amplification steps, thus optimizing sequencing time and efficiency. Additionally, the portability and affordability of the Nanopore sequencer MinION and Flongle, hold great promise to impact the clinical field. However, a major limitation has been the lack of analytical software featuring standardized parameters to aid in translation into clinical diagnostics.

Here we report our success in developing an amplification-free CRISPR-Cas9 targeted enrichment sequencing protocol using Nanopore MinION and Flongles to detect fusions relevant in the diagnosis and classification of CML and AMLs. The ONT Flongle is an adaptor for the MinION that provides cost-effective (∼$90 per flow cell), real-time sequencing for smaller assays. Our assays were designed to capture varied breakpoints of CML and APL, as well as fusion genes resulting from inv(16) (*MYH11-CBFB*) and t(4;11)(*KMT2A-AF4*). Simultaneous interrogation of these targets is a first step to a rapid characterization of AMLs in a single assay combining data that up to now required multiple different techniques and provide relevant information promptly. In addition to this amplification-free CRISPR-Cas9 nanopore assay, we extended our previously developed cloud-based nanopore data analysis pipeline ^13^ to include fusion detection and develop a custom breakpoint detection tool (see **Figure 1**). Using our optimized assay and our custom breakpoint finder, we showed that we can reliably detect and confirm fusion breakpoints in 80% of our specimens in under 3 hours of sequencing and data analysis.

**Figure 1.**
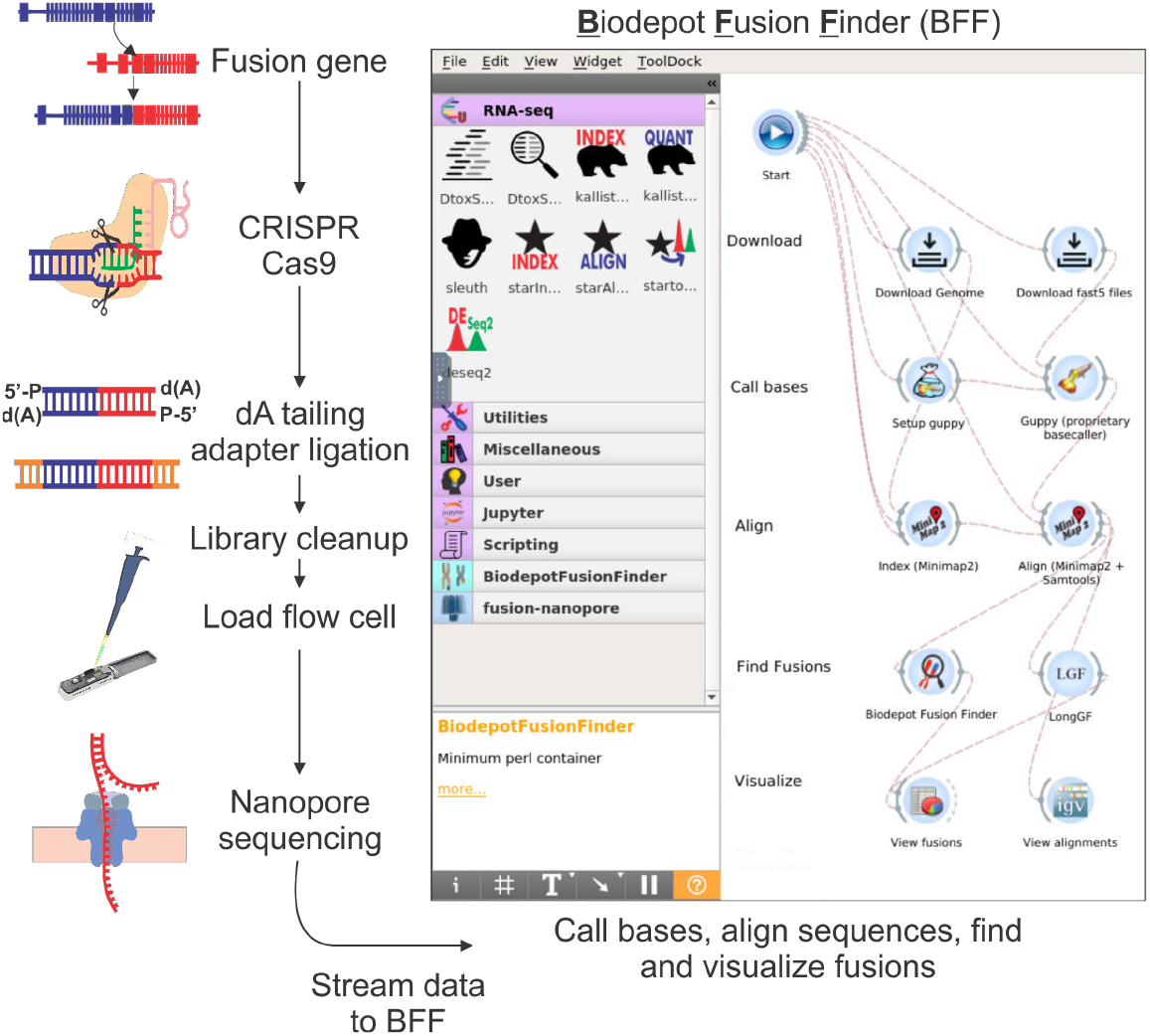
Chemistry and bioinformatics workflow of rapid single molecule long-read sequencing. Genomic DNA may contain a target fusion gene. The CRISPR-Cas9 system binds via specific guideRNA (gRNA) designed to enrich for DNA containing regions of interests. The library preparation does not undergo any amplification, and simply requires dA tailing and adapter ligation and a clean-up prior to being loaded onto a sequencing flow cell. On the flow cell libraries are sequenced by nanopores. Data from the sequencing devices are streamed onto biodepot workflow builder and are analyzed starting from FAST5 files, through an initial quality control, then base calling and alignment. After alignment different fusion finder tools were tested including LongGF and BFF before visualizing on IgV for confirmation and interpretation.

## Methods

### Cells lines and patient samples

Our assay was optimized using 6 cell lines: three with the *BCR-ABL1* fusion (K562, KU812, and KCL22), and NB4, MV4;11 and ME-1 that bear the *PML*-*RARA, KMT2A-AF4, and MYH11-CBFB* fusions respectively. Residual mononuclear cells from primary specimens (6 specimens from 5 patients with CML, 6 specimens from 5 patients with suspected APL, and 2 acute myeloid leukemia, not acute promyelocytic leukemia) were isolated using Ficoll® reagent (Millipore-Sigma) and banked in liquid nitrogen until the time of the experiment. All specimens had been originally tested in a CLIA certified laboratory according to standard clinical protocols.^14^ IRB coverage was obtained for use of residual laboratory samples. Patient samples were de-identified to the nanopore testing lab, and cytogenetic or molecular results were confirmed after nanopore results were rendered. Characteristics and demographics of specimens and patients are listed in **Table 1**.

**Table 1.** Characteristics of the Primary Specimens analyzed

| Patient Code | Age | Gender | Diagnosis | Specimen Source | Disease Burden | Cytogenetics/Molecular |
| --- | --- | --- | --- | --- | --- | --- |
| CML1 | 61 - 65 | Male | CML-AP Rlps | BM | 2% blasts | 46,XY,t(5;12)(q33;q15),t(9;22)(q34;q11.2)[2]/47,sl,+8[18] |
| CML2 | 41 - 45 | Female | CML-BC | PB | NA | Outside cytogenetics confirmed t(9;22) but only in 3/21 karyotypes. Full karyotype not available. |
| CML3 | 56 - 60 | Male | CML-AP | PB | NA | Patient with long standing p190 CML. At this time-point sample presents MDS/MPN with low level BCR-ABL1 by PCR. |
| CML4 | 21 - 25 | Male | CML-CP | PB | 3% blasts | 46,XX,t(9;22)(q34;q11.2)[20] |
| CML5 | 61 - 65 | Male | CML-AP Rlps | PB | 3% blasts | 46,XY,t(5;12)(q33;q15),t(9;22)(q34;q11.2)[2]/47,sl,+8[18] |
| CML6 | 65 - 70 | Female | CML-BC | PB | 16% blasts | 46,XX,t(9;22)(q34;q11.2)[4]; Confirmed by molecular studies. |
| AML1 | 36 - 40 | Male | AML | PB | 90% blasts | 46,XY,t(9;11)(q21;q23)[20] |
| AML2 | 36 - 40 | Female | AML | PB | 20% blasts | 47,XX+8,inv(16)(p13.1q22)[17]/46,XX[3] |
| APL1 | 81 - 85 | Male | APL | PB | <1% blasts | Patient with collision Multiple Myeloma and APL in BM with 30% blasts, not available for analysis. No circulating APL in PB. |
| APL2 | 31 - 35 | Female | APL | BM | 80% blasts | 46,XX,der(2)t(2;17)(q33;q21)t(15;17)(q22;q21),der(15)t(15;17),der(17)t(2;17)[3]/47,sl+8[3]/48,sdl,+8[12]/46,XX[2] |
| APL3 | 21 - 25 | Male | APL | PB | 75% blasts | NA |
| APL4 | 36 - 40 | Male | APL | BM | 83% blasts | AML with isochromosome 17q |
| APL5 | 36 - 40 | Male | APL | PB | 79% blasts | AML with isochromosome 17q |
| APL6 | 41 - 45 | Male | APL | BM | 22% blasts in a <5% marrow | 46, XY,t(1;7)(q21;q21), t(4;10)(q21;q25), add(8)(p23), del(9)(p22), t(11;17)(q23;q25)[4]/46, XY[16] from 3 months prior |

CML= Chronic Myeloid Leukemia; AML= Acute Myeloid Leukemia; APL= Acute Promyelocytic Leukemia, AP= Accelerated Phase, BC= Blast Crisis, PB= Peripheral Blood, BM= Bone Marrow; NA= not available

### Library preparation and sequencing assay

For the cell lines and 11/14 patient specimens the DNA was extracted with PureGene (Qiagen, Germantown, MD, USA) following the standard protocol. Special caution, including use of wide bore pipette tips and moderate centrifuge spin velocity was exercised to minimize fragmenting DNA strands. Two DNA specimens were extracted with AllPrep DNA/RNA Kit (Qiagen, Germantown, MD, USA), and one with QiAgen X-tractor with Reagent Pack DX (Qiagen, Germantown, MD, USA). cRNA guides were designed to direct Cas9 to cut on genomic proximity of each of the regions involved in each one of the translocations studied. When the target region was big, guides were tiled across the region to maximize coverage. Guides were designed to capture *PML-RARA, BCR-ABL1* p210, *KMT2A-AF4*, and *MYH11-CBFB*, including different fusion isoforms. Guides were designed using Chopchop (https://chopchop.cbu.uib.no/) with the CRISPR-Cas9 and nanopore enrichment settings and previously described ^15^.

We used 5 micrograms of DNA as input for each cell line and 2 to 5 micrograms for primary specimens. Average DNA integrity number (DIN) was 9.2 (range: 7.5-9.8). **Figure 1** shows a schematic of our workflow and details of the library prep are published^15^. Briefly, enrichment of target regions was obtained using Oxford Nanopore Technologies “Targeted, amplification-free DNA sequencing using CRISPR-Cas9” protocoll^12^. The different guides used in the assay were pooled in equimolar amounts of each guide. Through an initial dephosphorylation step, the 5’ ends of the DNA becomes inaccessible to adapter ligation. Double stranded DNA breaks that excise the region of interest are generated with the directional, target specific RNA guides complexed with tracrRNA and Cas9 enzyme. The Cas9 complex remains bound to the 5’ end of the guide, and the resulting new DNA ends contain a phosphorylated 5’end that is available for dA tailing and adapter ligation.^12^ All libraries generated in this manner were run on a MinION version 9.4. (Oxford Nanopore Technologies, Oxford, UK) nanopore sequencer using flow cells or Flongles. Modifications for libraries sequenced on the Flongle were only at the library loading step, in which the amount of Sequencing Buffer and library beads (both SQK-LSK 109, ONT) are reduced to from 35 to 13 and 25.5 to 7.5UL respectively, and 0.5UL of SQT is added. QC parameters tracked for each run are listed in **Table 2**.

**Table 2.**
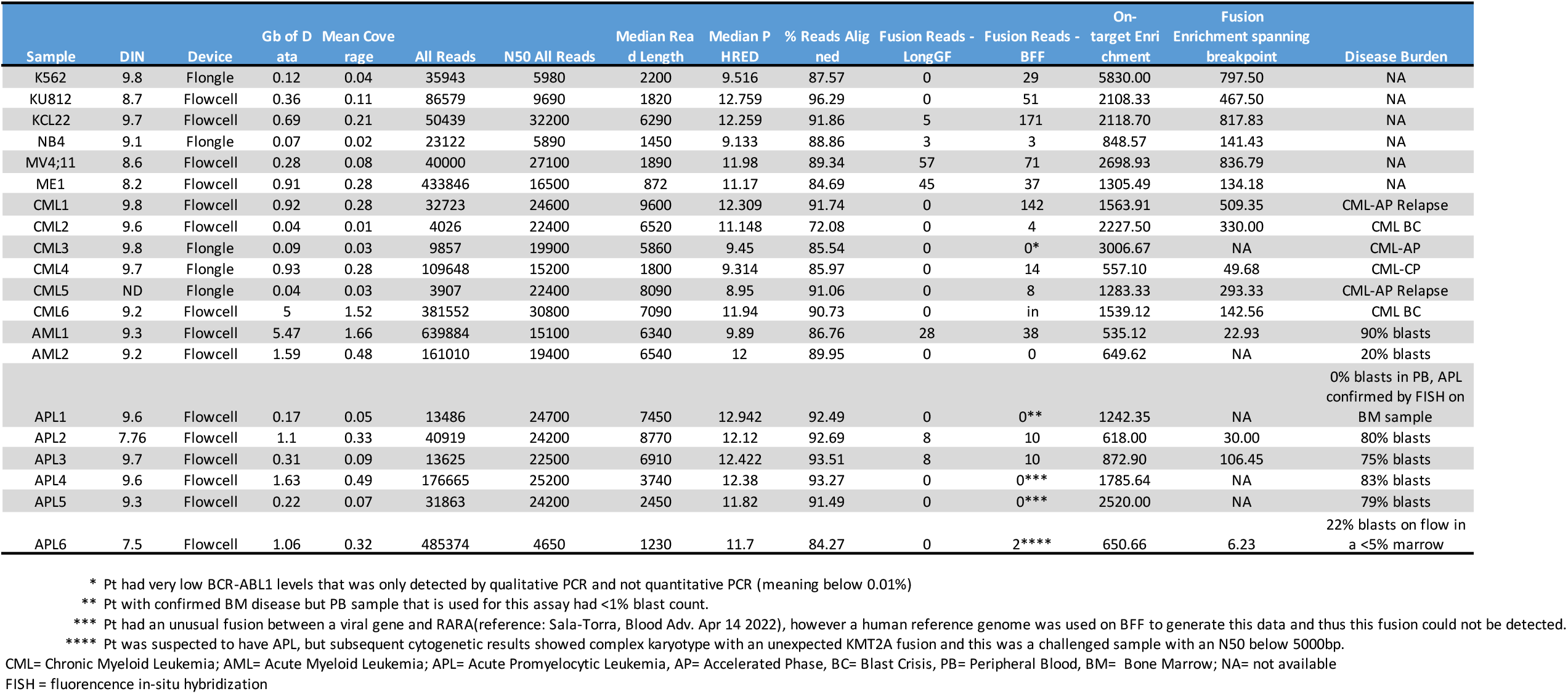
2. Characteristics of the Sequencing Runs

### PCR/Sanger sequencing

Primers specific to *BCR*-*ABL1* patient breakpoints were designed using Primer3 v. 0.4.0.^16^ in 2 cases. 100 ng of DNA were amplified, the PCR product was run on a 2% gel and Sanger sequenced to confirm the genomic breakpoint.

### Biodepot-workflow-builder: Interactive and accessible front end for fusion detection

We present a graphical, reproducible, and cloud-enabled fusion detection workflow consisting of all the steps of the analyses, including base calling, alignment, fusion detection, and visualization (see **Figure 1 screenshot of BwB interface)**. In contrast to the NanoFG workflow from Stangle et al^17^, our platform includes the computationally intensive base calling step, an interactive graphical user interface, and can readily leverage GPUs and be deployed on the cloud. Thus, analyses are fast, and our platform is accessible to biomedical and clinical scientists. Specifically, we extended the Biodepot-workflow-builder (Bwb)^18^ platform in which each computational task (or module) is represented by a graphical widget that calls a software container in the back end. Software containers, such as Docker, include all software dependencies and libraries required to execute the code. We have recently developed a Bwb workflow^13^ to support the processing of Nanopore data that includes the use of base callers Guppy^19^ and Bonito^19^, alignment using minimap2 and visualization of results using the Integrative Genomics Viewer (IGV) and GRCh37 hg19 as reference genome QC data was generated with PycoQC^20^. Guppy was used as the base caller. Minimap2^21^ was used as the aligner and variant caller. Fusions are visualized on IGV^22^ and confirmed on Blast. Most importantly, we extend our previous work by adding support for fusion detection, including LongGF^23^ and our own custom software “Biodepot Fusion Finder” (BFF).

### Bioinformatics pipeline for fusion detection: LongGF

LongGF^23^ is a software tool for fusion detection optimized for the high base calling error rates and alignment errors commonly found in long read sequencing data. LongGF takes as input a BAM file containing alignments (generated by minimap2 in this pipeline) and a GTF file containing the definitions of known genes. The output is a log file with detected gene fusions and their supporting reads. We created a graphical widget for LongGF in the Bwb. **Figure 2** shows a screenshot of the comparative workflows with output for LongGF versus BFF.

**Figure 2.**
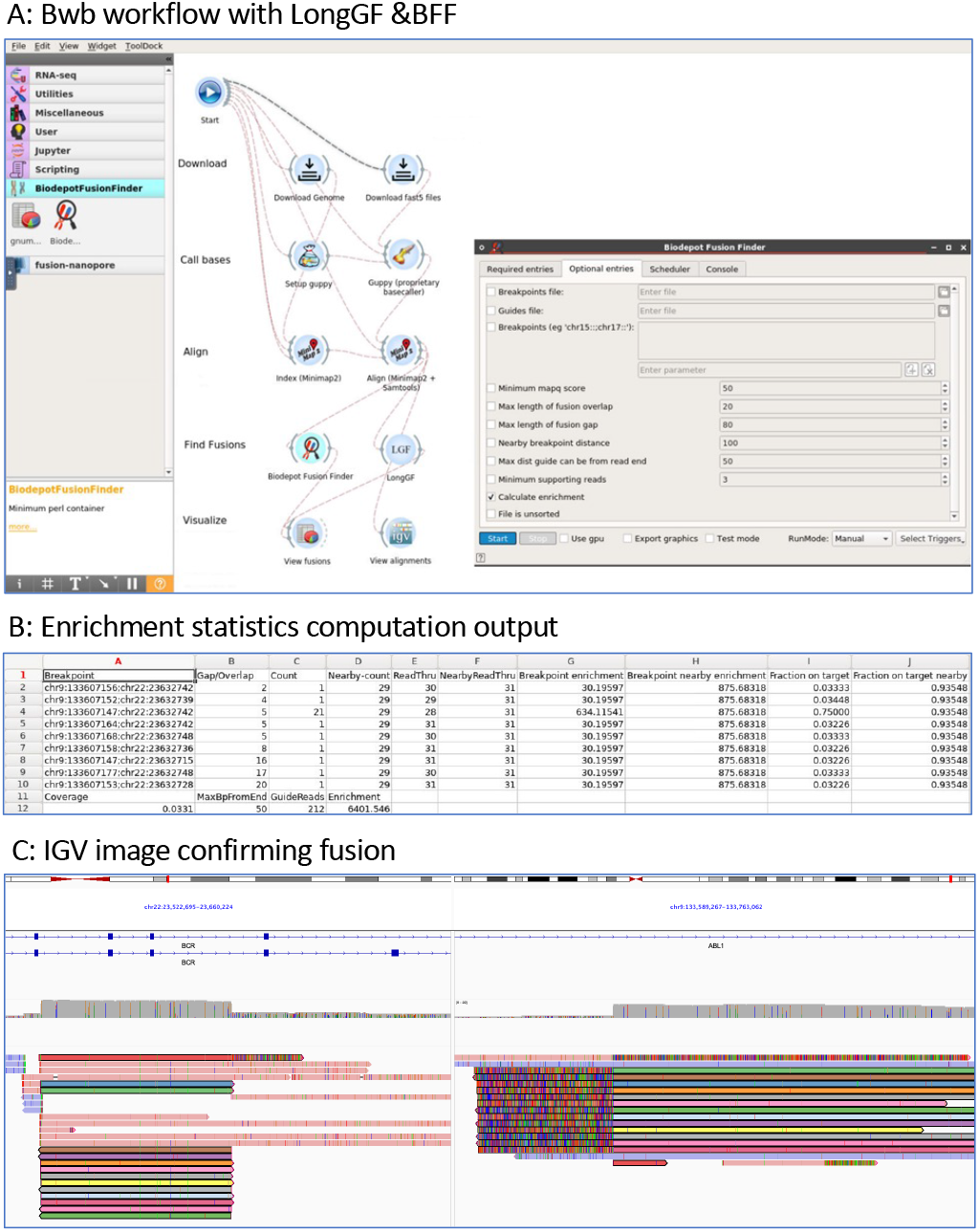
Screenshot of our bioinformatics workflow and output. **Panel A:** Bwb workflow showing the workflow including our custom Biodepot Fusion Finder (BFF) and LongGF widgets. **Panel B:** shows enrichment statistics as fusion enrichment and on target enrichment. **Panel C:** alignment can be viewed on IGV based on a BAM file generated from minimap2. This case shows a t(9;22) *BCR-ABL1* fusion in a primary specimen. Reads spanning the breakpoint are colored with the same color on alignment to both genes.

### Custom fusion detection tool: Biodepot Fusion Finder (BFF)

Reads that span a fusion gene will align to coordinates in both parts of the fusion and provide a specific breakpoint coordinate. We wrote a new software tool, the Biodepot Fusion Finder, BFF that examines the alternate alignments for each read and identifies reads that map to coordinates spanning a set of known breakpoints. This is similar to the strategy used by LongGF except that we allow for errors in the alignment near the breakpoint. Accordingly, the Breakpoint Finder identifies fusions that are not detected by LongGF which looks for supporting reads where the breakpoints are exactly matched. The user can provide a panel of breakpoint coordinates of interest. If no panel is provided, BFF will return candidate fusions that span nonadjacent genomic regions. Incomplete coordinates for breakpoints in the panel are supported – the user does not need to define the exact coordinates, nor do both breakpoints need to be given. BFF will return all the reads that match the panel of breakpoints in a text file for further inspection by the user if desired. The user can also provide a file with guide coordinates to obtain additional enrichment metrics. A containerized widget was developed that can integrated with the Bwb workflows for processing nanopore data. Using these workflows, we can directly analyze raw nanopore data and obtain lists of candidate fusions.

### Benchmarking experiments

We performed empirical experiments to benchmark the sensitivity and runtime required to reliably detect fusion. For each individual sample, sequencing metrics including quality scores and timestamps are obtained from the sequencing summary text file obtained as an output of base calling using Guppy. Detected fusions are then acquired from the Breakpoint Finder as well as LongGF. Specific fusion breakpoints are also provided in the data set from the BFF. All samples are then combined into a dictionary and separated by patient and cell line data. Plots are constructed for each category of data pertaining to the time to reach 3 fusions as well as the number of reads required to reach 3 fusions. Finally, the total number of fusion reads detected for each sample is compared between the BFF and LongGF as shown in **Figure 2**.

### Enrichment assessment

Two enrichment metrics were computed by BFF and tracked for each sample. First is the *fusion-specific enrichment* which is calculated with the following formula [(number of fusion reads) / (mean coverage of the genome)]. Second is the *on-target enrichment* which is calculated with the following formula [(number of reads that originate from a guide RNA cut point that includes the region of the breakpoint)/(mean genome coverage)]. Reads originating from a guide RNA cut are distinguished by the guide sequence being at the beginning of a read. As initial electrical signal data is generated by DNA passing through the nanopores (reads at the beginning of a strand) are error prone, which affects base calling and therefore the alignment of the reads so that the start sequence is often misaligned. Consequently, the BFF considers base pairs of the sequence near the start (default within 50 bp) of a read that aligns near the coordinates of a guide RNA cut site to have originated from a guide RNA cut. Specific reads cut by guides are manually confirmed. The allowed error intervals are customizable. SamTools v1.13 is used to sort and convert BAM files and determine the overall average coverage^24^. Picard CollectHsMetrics is used to generate the unique base pairs mapped metric for each sample^25^.

## Results

### Sample sequencing and enrichment

Details of the sample sequencing and enrichment metrics are included in Table 2. A range of 0.04 Gb – 5.47 gigabases of sequencing data was generated for each sample for an average mean coverage of the human genome of 0.32-fold (range: 0.01 – 1.66). We adopted standard quality metrics for nanopore workflows and tracked N50, which is a quality metric where half the reads are above this length (range 4.65kb-32.2kb) and median read length for total reads (range 0.87kb-9.60kb). Percentage of reads aligned ranged from 72 - 96%. Fusion specific enrichment was 135 - 837 fold for cell lines and 6 - 509 fold for patient samples. On target enrichment was 849 - 5830 fold for cell lines and 535 - 3007 fold for patient samples.

### Concordance of fusion detection with clinical results

Concordance with expected results was 100% for cell lines as the expected fusion was detected in 6/6 cell lines (3 *BCR*-*ABL1*, 1 *PML-RARA*, 1 *MYH11-CBFB*, 1 *KMT2A-AFF1*). Breakpoint sequences detected for *BCR-ABL1* cell lines are the same as previously published ^26^. We correctly confirmed the presence or absence of fusions in 11/14 (78.5%) primary specimens including both diagnostic and measurable residual disease (MRD) cases with a minimum of three reads, however one case (APL6) showed only 2 fusion reads and was not counted as confirmed. The 3 missed cases (CML3, APL1, and APL6 in tables 1 and 2) were comprised of low disease burden, ∼0%, 1% and <5%. We detected *BCR-ABL1* in 5/6 cases, *PML-RARA* in 2/6 cases, *KMT2A-AF9* and *CBFB-MYH11* in 1/1 case each. *BCR-ABL1* genomic breakpoint was confirmed by Sanger sequencing in CML1 and CML2 after designing patient specific primers.

In 3 specimens from 2 patients with suspected APL, we could not detect *PML-RARA* but observed other findings. Clinical and laboratory details are listed in Table 2. For the first patient, two specimens, one bone marrow and one peripheral blood (APL4 and APL 5) yielded no *PML-RARA* fusion reads. This patient presented with an AML morphologically suggestive of APL and an isochromosome 17q without t(15;17) detected by karyotype. While fusion detection software did not detect a fusion, manual inspection showed an insertion in an intronic region of RARA with TTMV viral genome; this case was previously reported^15^. Patient APL6 (presented with 22% blasts on flow in a <5% marrow which on unblinding showed a complex karyotype with t(11;17)(q23;q25) including the *KMT2A* gene. Two reads with *KMT2A-SEPT9* fusion were detected in a suboptimal but acceptable run (N50 < 5000bp; on-target enrichment 650.66 fold), confirming the lack of t(15;17) or *PML-RARA* fusion but the threshold was below the requisite 3 reads to confirm the *KMT2A-SEPT9* fusion.

### Comparison of fusion detection tools

A comparison of the bioinformatic workflows for data analysis using different fusion detection widgets including LongGF vs Biodepot Fusion Finder (BFF) was conducted; the specific workflow is demonstrated in **Figure 2**. Additionally, BFF computes the fusion enrichment and on target enrichment statistics; these are summarized in **Table 2**. In most cell line and primary specimens, LongGF shows a particular challenge in the detection of BCR-ABL1 and does not detect all fusion reads that are identified by BFF. Using the BFF, the average sequencing, and data processing time to 3 reads with fusions in the cell line experiments was 42.75 minutes (range: 18.87 - 77.65 min) and 188 minutes in the primary specimens where 3 reads were detected (range: 32 – 654 min) [see **Figure 3**, and **Supplemental Table 1**]. Cell line experiments took an average of 11,711 reads to identify 3 fusion reads (range: 1,321 – 43,326 reads) and 10,273 reads in primary specimens (range: 1,790 - 24,999 reads) for confirmation of the fusion calls.

**Figure 3.**
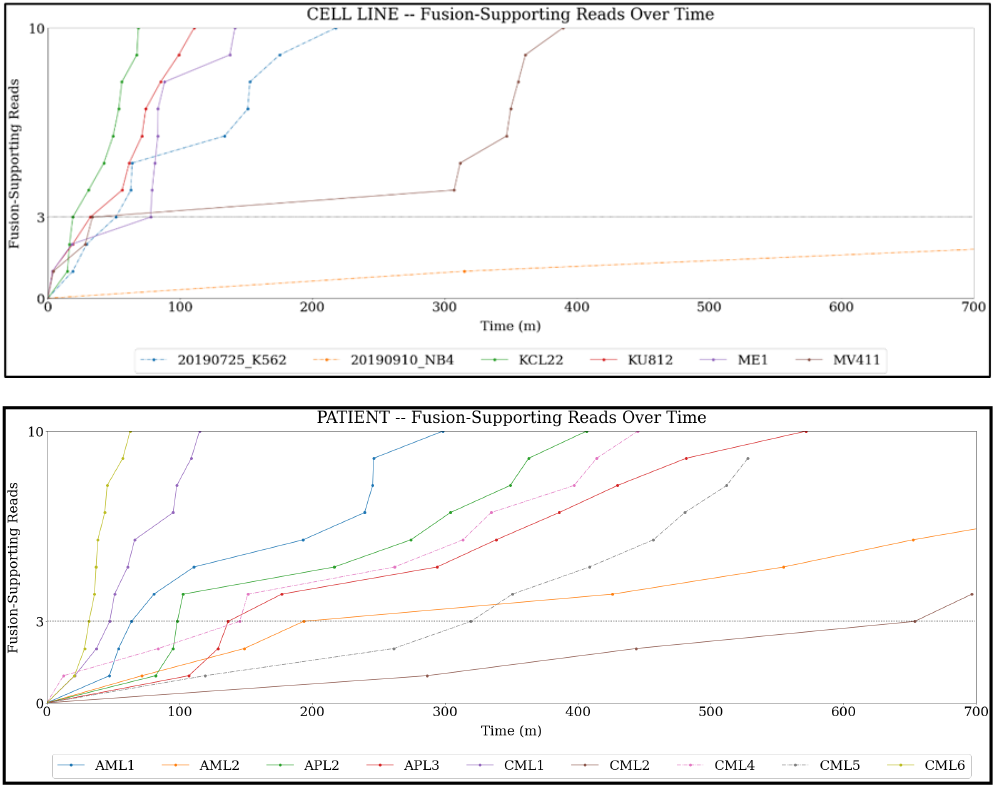
Time required to obtain 3 fusion supporting reads. Top panel shows the time to 3 fusion supporting reads seen in the cell lines samples. Bottom panel shows time to 3 fusion supporting reads seen in patient samples. Details of specific times are listed in supplementary table 1.

### Comparison of Flongles and flow cells

In five of our experiments (2 cell lines, 3 primary specimens), we used Flongles, while in the rest we used Flow Cells. The performance of the affordable Flongles was inferior to the Flow Cells, with lower expected average data output from the Flongles (based on manufacture expectations of ∼3GB), the N50 and median read length were smaller in Flongle reads (21,614 vs 19,166 and 6058 vs 5250 respectively), and significantly, the median Phred score for Flongle reads being lower than that for the Flow Cells (9.2 vs 11.88). Despite the worse performance of the Flongles, fusions were detected in 2 of 2 cell lines, and 2 of the 3 experiments with primary specimens with at least 8 and 14 reads confirming the fusion and the specimen without fusion confirmation was CML3 with pancytopenia and low disease burden.

## Discussion

We developed a rapid assay to detect fusion genes in blood or marrow samples in less than 8 hours with the fastest time achieving 3 fusion reads was 5 hours. This was accomplished by combining CRISPR-Cas9 enrichment during library preparation, nanopore long-read sequencing, a cloud-based data analytic pipeline, with a novel BFF program developed and optimized for finding fusions reads and breakpoints. RNA guides were designed to target genes involved in recurrent fusions in myeloid malignancies and used to enrich an amplification-free library preparation over 1600-fold. Our modular and containerized pipeline in Bwb allows users to efficiently process raw FAST5 data on the cloud through an accessible graphical user interface allowing for a very fast analysis step (average ∼4.5 minutes for basecalling, alignment, and fusion detection). To improve fusion calling, we developed the custom BFF that allow users to identify fusion reads not detected by LongGF. We successfully confirmed published genomic breakpoints in our series of cell lines and archival patient samples, which includes both diagnostic and follow-up samples to test feasibility in confirming both common and novel fusions over a range of tumor burdens. Our study includes 14 patient specimens and demonstrates the usability of this method in primary specimens with 2microgram of DNA.

An advantage of the CRISRP-Cas9 based enrichment protocol is that it allows for targeting of multiple common leukemia fusion genes by pooling multiple guide RNAs. Fusions are particularly well suited for this method as they have large gene segments that aid in alignment despite sequencing errors and wide variation where translocation breakpoints may occur. Other labs have employed similar but different methods that detected fusions by targeting one partner gene in the fusion ^17^. In contrast, our assay targets both partners of the fusion thus our approach allows for an expanded capability to detect known and novel fusions, such as in case AML1 with t(9;11), when guides are designed to target t(4;11). However, in 2/14 patients the assay did not detect fusion genes because both cases had a very low amount of fusion target. In one case (CML3) because the BCR-ABL1 was <0.01%, the other in the context of a very hypocellular sample.

Our work differs from standard RNA-based fusion detection assays, and instead interrogated single molecules of DNA rapidly and accurately to detect specific translocation breakpoints. Long-read sequencing technologies, like nanopore (Oxford Nanopore Technologies, Oxford, U.K.), allow sequencing of unamplified, long unbroken fragments of DNA which are more likely to span a breakpoint. This has potential clinical utility for personalized disease monitoring when CML patients are on TKI therapy and suppressing RNA transcription^27^; targeting DNA as a monitoring target may be more robust and reproducible since DNA is stable and present in constant numbers^28^. However genomic breakpoints in *BCR-ABL1* are unique to individual patients requiring patient-specific breakpoint characterization^28-32^ as *ABL1* breakpoints occur over an expansive region of about 150Kb, making this is an arduous endeavor previously involving multiple primer sets and Sanger sequencing^33^. Our method allows a single approach spanning the *BCR* and *ABL1* breakpoint regions without the use of multiple primers and PCR reactions. The sensitivity of DNA-based qPCR once the breakpoint is known can be as low as 10^−7 34-35^. Specimens CML1 and CML5 are a BM and PB obtained from the same patient and demonstrate high fidelity in confirming genomic breakpoints and the ability to use patient specific primers for personalized MRD monitoring.

Advantages of long-read sequencing over current clinical diagnostic assays are speed and the relatively low complexity of the assay when compared to cytogenetics and targeted NGS panels. While long-read sequencing results could potentially have a turnaround time (TAT) of less than 24 hours, full karyotype analysis TAT is generally longer with the fastest times at days to a week and most targeted NGS panels require ∼7-10 days from start of processing to result report. The nanopore sequencing streams data simultaneously to our GPU-enabled data analytic pipeline in the Bwb interface which resides on the cloud to help interpret and reliably identify fusion reads within 5000 seconds(<2 hours) computational time in most specimens. Building on our experience^14,17^, we used 3 reads as a threshold for fusion confirmations. With current simultaneous sequencing and data analysis workflow described here, 3 sequences are detected in an average of 3 hours and 7 min (fastest at 30 min) in the 9 patient specimens where a fusion could be confirmed. This means that a diagnostic result with a precise fusion breakpoint with 3 fusion supporting reads would be possible in the same day.

Five specimens were sequenced on the less costly Flongle device, which has lower sequencing capabilities (pore count ∼60, sequencing life 24h), but 3 fusion reads were reached in 4/5 specimens (2 cell lines, 3 patient samples) with less resources. The sequencing quality is significantly lower on the Flongles (median Phred 9.2 in Flongles vs 11.9 in Flow Cells), however fusions were detected in all samples with adequate tumor burden. To achieve a cheaper version of our fast and portable nanopore fusion assay, further challenges will need to be addressed. We predict overall less sequencing can be achieved, even with additional optimizations of the CRISPR library guides as multiplexing of the guide RNA appears to increase efficiency and on-target fusion reads. Nanopores on the Flongles have poorer viability and generate more read errors demonstrable by lower Flongle Phred quality on average and therefore additional alignment challenges and a study of errors specific in Flongle bioinformatic data compared to the flow cells are needed to understand potential compensation mechanisms for data analysis.

## Conclusion

We demonstrate the feasibility of using single molecule long-range sequencing assay to detect fusion genes in heme malignancy (AML, CML and APL) patients. Inherent characteristics of fusions make this assay a promising cost effective tool for rapid detection of recurrent fusions that 1) does not require previous knowledge of the target,, 2) with a rapid TAT (8 hours in 80% of samples) when multiplexing different assays and used with the specific data analysis and fusion detection tools described in our manuscript, 3) can precisely map translocation genomic breakpoints that allow for development of personalized markers for disease monitoring, and 4) can potentially allow discovery of novel/different fusion partners

## Data Availability

Data from this study are available upon request to the authors.

## Contributions

OST and CY designed the study and performed background research and secured IRB protocol. KYY and LHH designed the informatics pipeline and algorithms. JR, DW and CY identified specimens. LHH and SR implemented the software. SR, OST and CY tested the software. OST, LB, SR, LHH, CY performed research and analyzed the data. KYY, JR, CY funded the project and provided supervision. OST, SR, KYY, and CY drafted the manuscript. All authors reviewed and edited the manuscript.

## Acknowledgements

The authors want to thank Dr Phillip E. Starshak, Kaiser Permanente Oakland, for procuring specimens.

## Figure and Table legends

**Supplementary Table 1**. Times and total reads before 3 fusion reads could be confirmed in cell lines and patient samples.

