## Supplemental Table 1 for "Rapid detection of myeloid neoplasm fusions using Single Molecule Long-Read Sequencing"

Supplementary Table 1. Times and total reads before 3 fusion reads could be confirmed in cell lines and patient samples.

| Cell lines | Time to 3 Fusions (min) | Reads to 3 Fusions |
|---|---|---|
| K562 | 51.44 | 2598 |
| KCL22 | 18.87 | 1321 |
| KU812 | 32.16 | 4137 |
| ME1 | 77.65 | 43326 |
| MV411 | 33.65 | 7175 |
| NB4 | 831.26 | 22803 |
| **Patient samples** | **Time to 3 Fusions (min)** | **Reads to 3 Fusions** |
| CML1 | 47.33 | 2093 |
| CML2 | 653.56 | 2869 |
| CML3 | NR | NR |
| CML4 | 145.23 | 21201 |
| CML5 | 319.42 | 1790 |
| CML6 | 31.74 | 10303 |
| AML1 | 63.78 | 17414 |
| AML2 | 193.36 | 24999 |
| APL1 | NR | NR |
| APL2 | 98.27 | 9106 |
| APL3 | 136.58 | 2683 |
| APL4 | NR | NR |
| APL5 | NR | NR |
| APL6 | NR | NR |

min= minutes, NR= Not reached
